# Association between enterotypes of the gut microbiota and features of stroke

**DOI:** 10.1101/2024.05.15.24307460

**Authors:** Toshiyasu Ogata, Hisatomi Arima, Miki Kawazoe, Yasuhiko Baba

**Affiliations:** Department of Neurology, Faculty of Medicine, Fukuoka University, Fukuoka, Japan; Department of Neurology, Japanese Red Cross Fukuoka Hospital, Fukuoka, Japan; Department of Preventive Medicine and Public Health, Faculty of Medicine, Fukuoka University, Fukuoka, Japan

**Author notes:** Corresponding author and reprint requests to: Toshiyasu Ogata, MD, PhD Department of Neurology, Japanese Red Cross Fukuoka Hospital, 3-1-1, Okusu, Minami-ku, Fukuoka 815-8555, Japan.

**Keywords:** ischemic stroke, gut microbiota, atherosclerosi

## Abstract

**Background:** An enterotype (e.g., genera *Bacteroides* and *Prevotella*) is a classification of patients’ gut microbes into three types, and these types differ in their features of cardiovascular disease. We hypothesized that patients have different enterotypes depending on their arteriosclerosis risk factors, stroke subtype, and severity of stroke.

**Methods:** Stool specimens were collected from 100 patients (age: 73.4 ± 11.3 years, 62 men, 38 women) with ischemic stroke after consent was obtained. Data on age, sex, risk of arteriosclerosis, stroke subtype, history of stroke, neurological severity at admission, and prognosis were obtained from the patients’ medical records. Phylogenetic analyses of the 16S rRNA gene (V3–V4 region) extracted from each stool sample were performed. Quantitative analyses of the presence of each bacterial genus in the intestines were performed using a next-generation sequencer. After the number of each genus of gut microbes was extracted, ≥ 30% of the patients with the genus *Bacteroides* were classified as type I, ≥ 15% with the genus *Prevotella* were classified as type II, and the rest were classified as type III. We analyzed the association between the patients’ enterotypes and their characteristics (i.e., arteriosclerosis risk factors such as stroke subtype, and severity of stroke).

**Results:** Thirty-three patients had type I, 10 had type II, and 57 had type III, with no overlap. Patients with types I and II had a lower prevalence of dyslipidemia than those with type III (types I vs II vs III: 36% vs 20% vs 58%, P = 0.028), a lower National Institute of Health and Stroke Scale score at admission (1 vs 1 vs 4 [median], P = 0.025), and the modified Rankin Scale score at discharge tended to be lower (1 vs 1 vs 2 [median], P = 0.094).

**Conclusions:** The enterotype may affect the risk factors and severity of ischemic stroke.

---

The gut microbiota has been increasingly attracting attention as the major cause of various diseases because of the recent development of metagenomics. Atherosclerosis is strongly associated with diet. Therefore, the gut microbiota has long been suspected to play a crucial role in the progression of atherosclerosis.

In the gut microbiota, the genus *Bacteroides* has attracted much research focus because it is frequently found in feces. The association of *Bacteroides* with a high-fat diet has been reported mainly in Western countries^1^. One study reported that people with a high level of *Bacteroide*s in their fecal samples had a diet high in protein and fat^2^. Another study showed that the genera *Bacteroides, Alistipes*, and *Bilophila* were increased in people who ingested a diet rich in animal fat for several days^3^. However, people with a high body mass index were reported to have a lower rate of *Bacteroides* than those with a low body mass index^4^.

In contrast, some researchers from East Asia disagree with the opinion that the genus *Bacteroides* may detrimentally affect atherosclerosis. A study from Japan showed that the abundance of the phylum Bacteroidetes was decreased and that of the phyla Firmicutes and Actinobacteria were increased in patients with type 2 diabetes compared with healthy subjects^5^. A Chinese study showed that the abundance of *Bacteroides* and *Prevotella* was decreased in patients with symptomatic atherosclerotic ischemic stroke and transient ischemic attack^6^.

Arumugam et al suggested that the human gut microbiota could be stratified into three enterotypes, which could help identify the characteristics of people and their diseases^7^. Yamashita et al found that, using this classification, patients with coronary heart disease had a low rate of *Bacteroides* (< 30%)^8^. This study suggests the usefulness of the enterotype classification.

Although several studies have reported on the gut microbiota in patients with ischemic stroke^6, 9^, there have been few reports on the association between enterotypes and atherosclerosis in patients with stroke, especially regarding the association of *Bacteroides8*. Therefore, this study aimed to determine the association of enterotypes with the features of stroke, especially the progression of atherosclerosis.

## Methods

We prospectively registered 100 patients (73.4 ± 11.3 years; 62 men, 38 women) who were hospitalized for the treatment of ischemic stroke between October 2021 and December 2022. All of the patients were admitted to our neurological center within 7 days following the occurrence of ischemic stroke, and they were then informed about and consented to participate in this study. We diagnosed patients with ischemic stroke when they had a rapid onset of focal brain disturbance. We confirmed the findings with diffusion-weighted imaging and conducted carotid ultrasound for the evaluation of carotid stenosis. We examined the patients’ medical records to identify stroke risk factors, such as age, sex, stroke subtype, hypertension, diabetes mellitus, hyperlipidemia, atrial fibrillation, current smoking and drinking habits, and a previous history of stroke and cancer. We also assessed their medical data to collect information on the National Institute of Health and Stroke Scale (NIHSS) score, stroke subtype, and outcome at discharge. The stroke subtype was classified using the TOAST criteria^10^. Furthermore, SVO_LAA was defined as small vessel occlusion (SVO) or large artery atherosclerosis (LAA) because these are the stroke subtypes related to atherosclerosis. The outcome at discharge was evaluated by the modified Rankin Scale; a score of 0–1 was considered favorable because the patients’ condition was not severe. This study was approved by the ethics committee of Fukuoka University Hospital (IRB no.: H20-08-008). We adhere to the observational cohort guideline of STROBE.

We extracted data from carotid ultrasound for all of the patients. We collected the maximum intima–media thickness of each side of the common carotid artery and calculated their average (average value of the maximum common carotid intima–media thickness), and the presence of plaque and stenosis ≥ 50% on the carotid bifurcation on either side. An atheromatous plaque was defined as a lesion with an intima–media thickness ≥ 1.1 mm.

### Gut microbial measurements

A brush-type fecal collection kit (TechnoSuruga Laboratory Co., Ltd., Shizuoka, Japan) containing preservation fluid was used to obtain and stably maintain the microbiota in feces, and was preserved in the refrigerator for up to 3 weeks. The kits were then sent to Bioengineering Lab. Co., Ltd. (Sagamihara, Japan) to extract and analyze DNA from fecal fluid using the bead-beating method, and to perform 16S rRNA gene sequencing.

DNA extracts were used as templates to amplify the V3–V4 region of each 16S rRNA gene with the primer pair 341F/805R using the two-step tailing polymerase chain reaction method. Paired-end sequencing of polymerase chain reaction amplicons was performed on the Illumina MiSeq platform (Illumina Inc., San Diego, CA) at 2 × 300 base pairs.

### Statistical analysis

The patients were divided into three groups as follows: patients with a rate of *Bacteroides* ≥ 30% (enterotype I group); those with a rate of *Prevotella* ≥ 15% (enterotype II); and those with genera other than enterotype I or II (enterotype III group).

The differences between the patients’ characteristics were statistically analyzed between the three groups. We also analyzed the differences in the patients’ stroke subtype, SVO_LAA, and the TOAST classification between the three groups. If the patients’ characteristics were significantly different among the three groups, age- and sex-adjusted analyses were also conducted. All data were analyzed using SPSS v28.0 software (IBM Corp., Armonk, NY).

## Results

There were 33 patients in the enterotype I group, 10 in the enterotype II group, and 57 in the enterotype III group. There was no overlap between those who were classified as groups I and II. The median time from stroke onset to the collection of stool specimens was 3 days. The median NIHSS score was 2. The percentages of hypertension, diabetes mellitus, dyslipidemia, atrial fibrillation, and smoking and drinking habits were 80%, 38%, 47%, 24%, 22%, and 24%, respectively. Regarding the stroke subtype, the percentages of SVO, LAA, and cardioembolic stroke were 17%, 33%, and 22%, respectively, while that of other stroke etiologies and undetermined causes was 28%. Regarding the gut microbiota of the patients, the mean percentages of *Bacteroides* and *Prevotella* were 24.7% ± 11.2% and 4.0% ± 8.9%, respectively.

The patients’ background and atherosclerotic risk in the three groups are shown in Table 1. There was a significant difference in the frequency of dyslipidemia (P = 0.028) between the three enterotype groups. The rate of diabetes mellitus appeared to be lower in the enterotype II group than in the other groups, but this was not significant. The NIHSS score at admission was significantly higher in the enterotype III group than in the other groups (P = 0.025). The modified Rankin Scale score at discharge tended to be worse in the enterotype IIIgroup than in the other groups (P = 0.094). The degree of atherosclerotic changes evaluated by carotid ultrasound was similar in the three groups.

**Table 1:** Patients’ characteristics depending on the enterotype.

|  | Enterotype I | Enterotype II | Enterotype III | p |
| --- | --- | --- | --- | --- |
|  | N=33 | N=10 | N=57 |  |
| Age | 70.6±12.3 | 76.3±11.5 | 74.5±10.6 | 0.21 |
| Gender (female) | 12(36.4%) | 4 (40%) | 22 (38.6%) | 0.97 |
| BMI | 21.7±3.1 | 21.9±3.7 | 22.6±2.7 | 0.37 |
| Hypertension | 25 (75.8%) | 9 (90%) | 46 (80.7%) | 0.6 |
| Diabetes mellitus | 13 (39.4%) | 1 (10%) | 24 (42.1%) | 0.15 |
| Dyslipidemia | 12 (36.4%) | 2 (20%) | 33 (57.9%) | 0.028 |
| Smoking habit | 7 (21.2%) | 2 (20%) | 7 (12.3%) | 0.5 |
| drinking | 6 (18.2%) | 3 (30%) | 13 (22.8%) | 0.71 |
| Atrial fibrillation | 6 (18.2%) | 3 (30%) | 15 (26.3%) | 0.61 |
| Ischemic heart disease | 4 (12.1%) | 1 (10%) | 7 (12.3%) | 0.98 |
| History of stroke | 5 (15.2%) | 1 (10%) | 18 (31.6%) | 0.12 |
| History of cancer | 3 (9.1%) | 0 (0%) | 3 (5.3%) | 0.206 |
| SVO_LAA | 12 (36.4%) | 6 (60%) | 32 (56.1%) | 0.16 |
| NIHSS at admission | 1 (0-5) | 1 (1-2) | 4 (1-6) | 0.025 |
| mRS at discharge | 1 (1-3) | 1 (0-2) | 2 (1-3) | 0.094 |
| IMT-C10 | 1.36±0.86 | 1.34±0.64 | 1.26±0.53 | 0.78 |
| maximum of max IMT | 2.43±1.13 | 2.44±1.20 | 2.41±1.18 | 0.99 |
| presence of plaque | 29 (87.9%) | 10 (100%) | 55 (96.5%) | 0.18 |
| area stenosis≥50% | 14 (42.4%) | 5 (50%) | 18 (31.6%) | 0.4 |

Enterotype I: group of patients with a rate of *Bacteroides* $\geq 30\%$ ; enterotype II: group of patients with a rate of *Prevotella* $\geq 15\%$ ; enterotype III: group of patients with a genus other than that in enterotype I or II.

Age- and sex-adjusted analyses are shown in Table 2. Using the enterotype III group as a reference, the odds ratio of diabetes mellitus was lower in the enterotype II group, but this was not significant. However, the odds ratios of dyslipidemia in the enterotype I and II groups were significantly lower than that in the enterotype IIIgroup. The odds ratio of the NIHSS score at admission was significantly lower in the enterotype II group than in the enterotype I group, while that of the enterotype I group was also low. The odds ratio of the modified Rankin Scale score at discharge was also significantly lower in the enterotype II group than in the enterotype I group. Among the microbes in the enterotype III group, the rates of the classes Lachnospiraceae and Ruminococcus in the phylum Firmicutes were increased (Table 3).

**Table 2:**
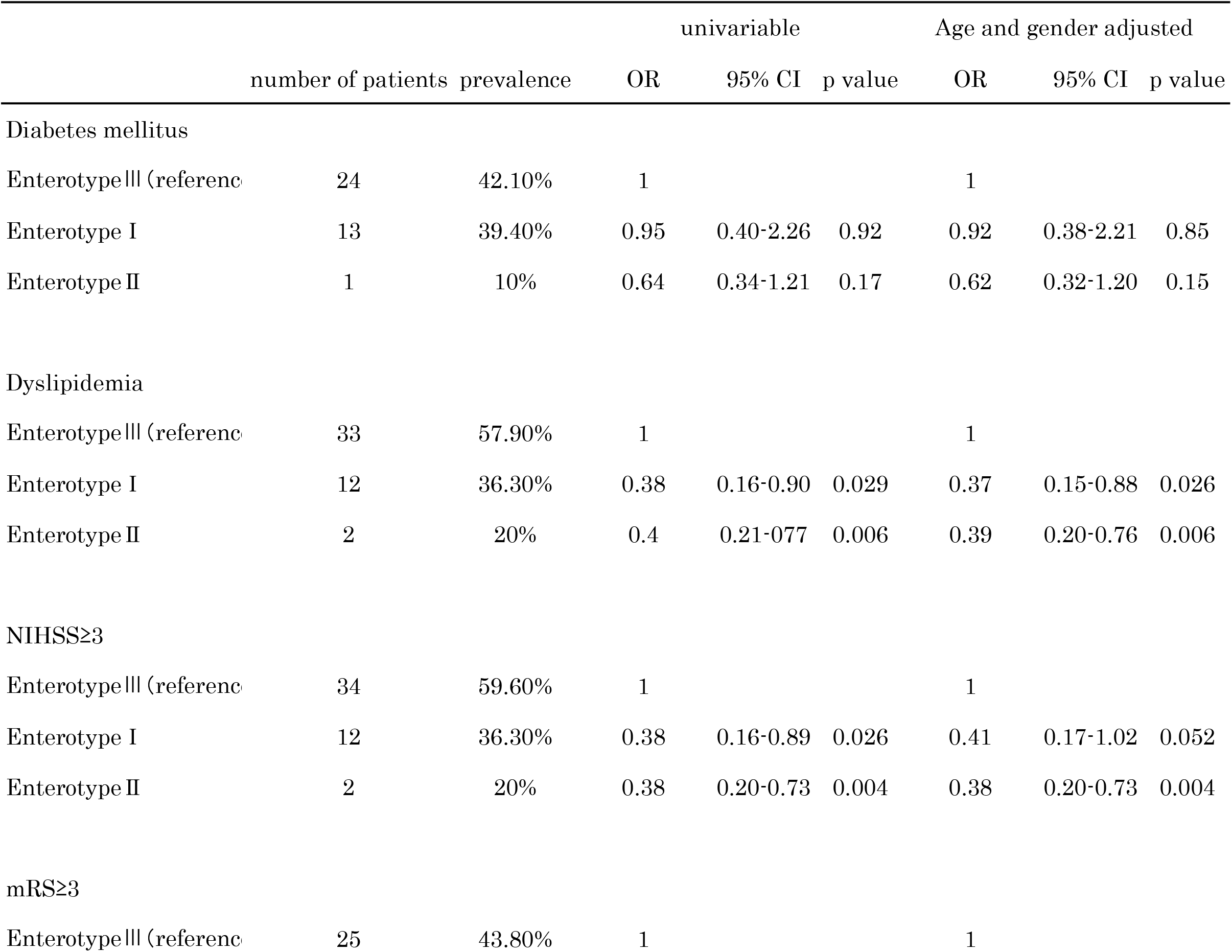
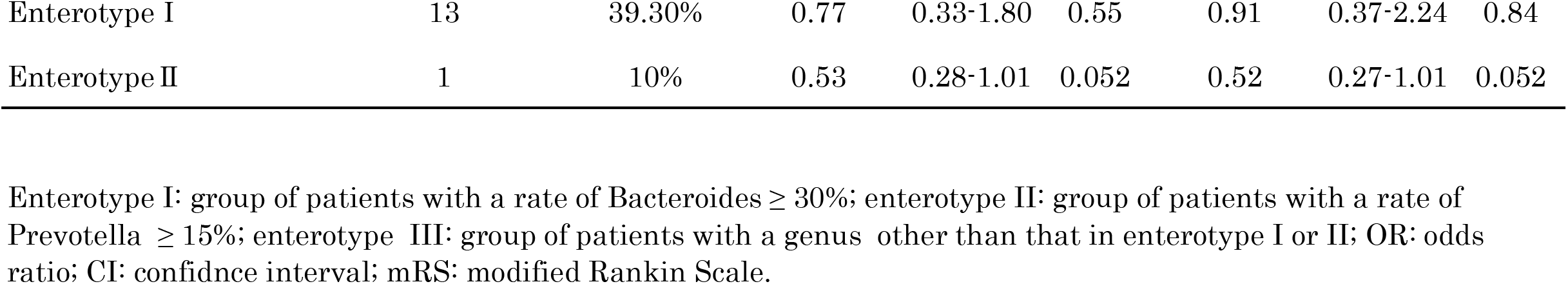
Age- and sex-adjusted analyses.

**Table 3:**
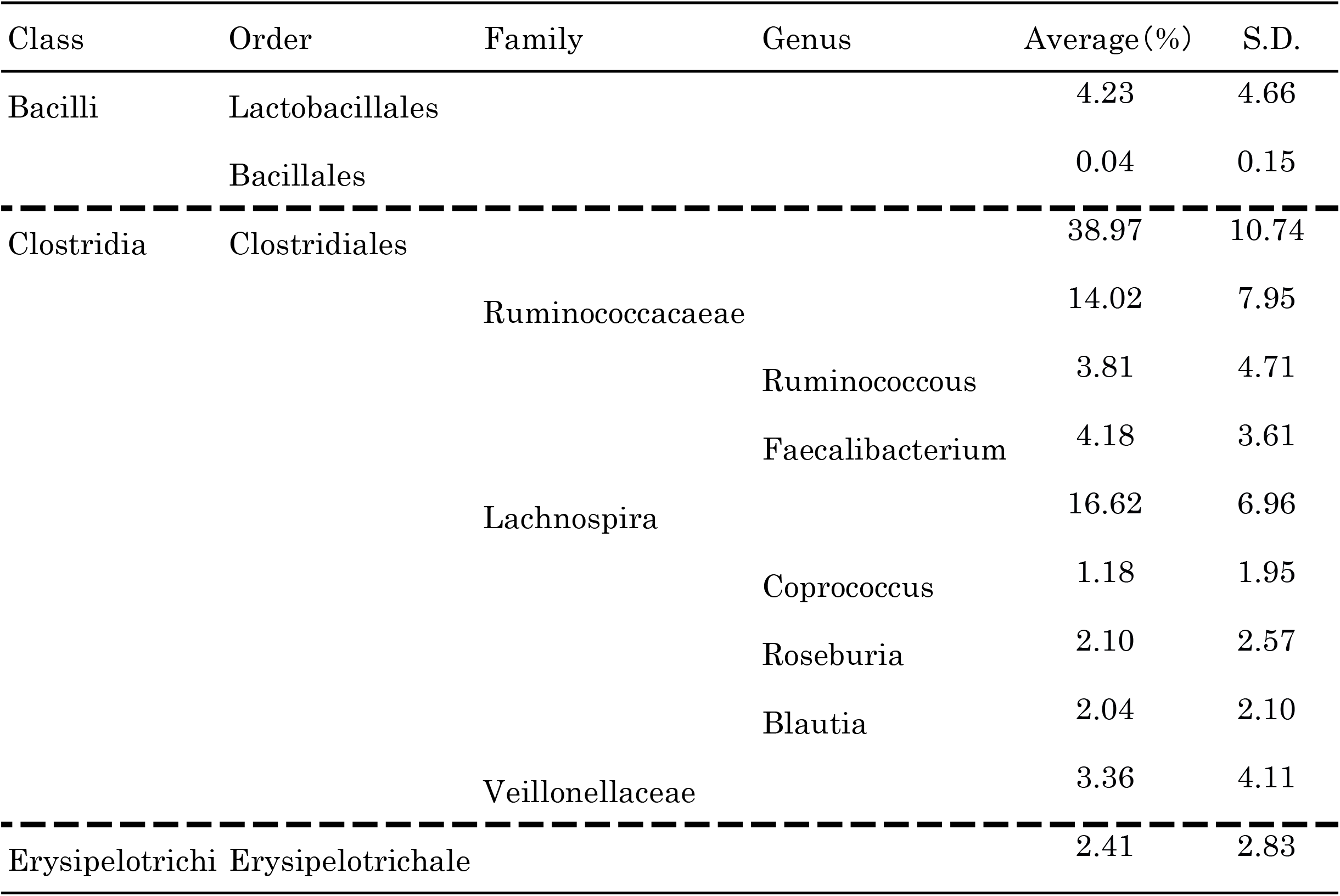
Main gut microbes in enterotype group III.

## Discussion

In this study, we classified patients with stroke into three enterotypes and examined their association with arteriosclerosis risk factors, stroke subtype, and severity of stroke. This study showed that the enterotype was significantly associated with the frequency of dyslipidemia and stroke severity.

The abundance of *Bacteroides* is decreased in patients with obesity^11, 12^ or diabetes^13^. However, a study showed that *Bacteroides* and *Prevotella* may be increased in participants who were more adherent to a Mediterranean diet^1^. The Mediterranean diet is characterized by a healthier diet with a higher intake of fiber, vegetables, and fruit, and with a lower intake of sugar and red meat. These findings indicate that characteristics and effects on the human body might vary depending on the region and culture. The classification used in the current study is the same as that from Japan, which indicates the usefulness of identifying atherosclerosis in this study. Therefore, *Bacteroides* might have beneficial roles in the prevention of arteriosclerosis in Japan.

In this study, the rates of Lachnospiraceae and Ruminococcus were increased in the enterotype III group. An increase in Ruminococcus has been reported to be associated with diabetes^14^. In contrast, Lachnospiraceae has been reported to be caused by an increase in non-meat meals and is associated with butyric acid^15^. The abundance of Firmicutes is increased in the gut flora of Japanese people with diabetes, which is different from the distribution of the gut microbiota in Western countries. There have been no reports on the role of Lachnospiraceae or Ruminococcus in hypertension, which is considered to be the greatest risk for brain infarction. Further studies are required to determine the role of enterotype 3 on brain infarction.

The precise mechanisms of *Bacteroides* and *Prevotella* on the prevention of atherosclerosis remain unknown. However, the abundance of *Prevotella* was significantly lower in fecal samples of Japanese patients with diabetes, while that of Lactobacillus was higher, than in control subjects^16^. In the current study, the rate of *Prevotella* appeared to be decreased with the presence of diabetes and hyperlipidemia. *Bacteroides* was found to have a beneficial role in glucose metabolism and was less abundant in patients with type II diabetes mellitus^17^. Another report showed that a lower abundance of *Bacteroides* and an increased Firmicutes to Bacteroidetes ratio were associated with the incidence of cardiovascular disease^18^. *Bacteroides* and *Prevotella* might contribute to the prevention of atherosclerosis. Therefore, further research is necessary to identify the role of enterotypes in the progression and suppression of inflammation of vessel walls.

The composition of the gut microbiota can be remarkably diverse, and there is no clear definition of a healthy gut microbiota. Inflammation of the vessel walls may lead to the progression of atherosclerosis. Therefore, clarifying whether the progression of arteriosclerosis can be suppressed by regulating the gut microbiota, such as increasing *Bacteroides* or *Prevotella*, is necessary.

There are several limitations to this study. First, the number of patients in this study was small. Second, this study analyzed enterotypes and did not address each species, which might be a limitation because the function of the gut microbiota could vary from species to species.

In conclusion, enterotypes may affect the risk factors and severity of ischemic stroke. An increase in abundance of *Bacteroides* and *Prevotella* could play a beneficial role in the prevention of atherosclerosis.

## Data Availability

The participants of this study did not give written consent for their data to be shared publicly, so due to the sensitive nature of the research supporting data is not available.

## Acknowledgments

We thank Ellen Knapp, PhD, from Edanz (https://jp.edanz.com/ac) for editing a draft of this manuscript.

## Non-standard Abbreviations and Acronyms

NIHSS: National Institute of Health and Stroke Scale
TOAST: Trials of Org 10172 in Acute Stroke Treatment
SVO: small vessel occlusion
LAA: large artery atherosclerosis

